# Oral Azvudine (FNC) Tablets in Patients infected with SARS-CoV-2 Omicron Variant: A Retrospective Cohort Study

**DOI:** 10.1101/2023.01.05.23284180

**Authors:** Wenmei Chen, Honggang Xu, Lei Hong, Ran Yang, Caiqiu Peng, Guiqiang Wang, Wei Li

**Affiliations:** Department of Respiratory Disease, the First Affiliated Hospital of Bengbu Medical College, Bengbu, 233004, China; Department of Infectious Diseases, Peking University First Hospital, Beijing 100034, China; Department of Orthopedics, the First Affiliated Hospital of Anhui Medical University, Hefei, 230022, China; Center for Clinical Medicine of Respiratory Disease(tumor), Bengbu, Anhui, 233004, China; Provincial Key Laboratory of Respiratory Disease, Bengbu, Anhui, 233004, China; Department of Infectious Diseases, Peking University International Hospital, Beijing, 102206, China; Beijing Key Laboratory of Hepatitis C and Immunotherapy for Liver Diseases, Beijing, 100034, China

**Keywords:** SARS-CoV-2, Omicron variant, Azvudine, Virus shedding time, Prognosis

## Abstract

**Background:** There is a lack of data on the efficacy of oral Azvudine in Coronavirus Disease treatment. This study aimed to assess the association between Azvudine treatment and clinical outcomes in a cohort of patients infected with the SARS-CoV-2 Omicron variant.

**Methods:** This is a retrospective study conducted in two mobile cabin hospitals. All consecutive patients with a diagnosis of COVID-19 admitted from August to October 2022 were included in the study. Linear regression models and Cox proportional hazards models were used to assess the association between Azvudine treatment and time to obtain the first negative nucleic acid test results.

**Results:** A total of 207 patients were analyzed, of whom 166 patients (80.2%) received Azvudine treatment after hospitalization, and the rest did not. Linear regression models showed that Azvudine treatment was associated with reduced time to obtain the first negative nucleic acid test results after adjusting for age and gender [mean difference = −1.658; 95% CI: −2.772 to −0.544, P = 0.0039]. The multivariable Cox analysis conforms to the results from the linear regression model (hazard ratio = 1.461; 95% CI: 1.01 to 2.11, P = 0.044).

**Conclusion:** Azvudine treatment was associated with reduced virus shedding time. Further studies are needed to confirm our findings.

## Introduction

The Coronavirus Disease (COVID-19) epidemic caused by severe acute respiratory syndrome coronavirus 2 (SARS-CoV-2) is spreading across the world [1, 2]. In November 2021, the Omicron variant (B.1.1.529) of SARS-CoV-2 emerged and quickly exceeded other variants of concerns (VOCs) and became the dominating one[3]. The high number of mutations in the spike protein promotes immune evasion [4], leading to enhanced transmission ability and weakened protective effects from neutralizing antibodies [5].

Antiviral drugs are considered a potential therapy and are recommended in guidelines [6, 7]. Azvudine (also known as FNC), a 4’-modified nucleoside drug candidate [8], is the first domestic-developed oral antiviral agent in China[9]. A pilot study compared Azvudine with standard antiviral treatment in 20 patients with mild and moderate COVID-19, and found that oral Azvudine tablets could shorten the viral shedding time [10]. However, this study has a limited number of patients and only focuses on mild and moderate COVID-19 patients.

Given the scarcity of data on the oral Azvudine tablets in patients infected with the SARS-CoV-2 Omicron variant, we conducted a retrospective study to assess the association between Azvudine treatment and clinical outcomes in a cohort of patients infected with SARS-CoV-2 Omicron variant.

## Methods

### Study design and patients

This study is a multicenter, retrospective study conducted in Luqiong and Naidong mobile cabin hospitals, which are novel concept hospitals for treating non-critical COVID-19 patients. This study was approved by the institutional review board of the First Affiliated Hospital of Bengbu Medical College with a waiver of consent for the the retrospective observational design (No. 2020KY112, 2020KY016). All consecutive patients with a diagnosis of COVID-19 admitted from August to October 2022 were included in the study. SARS-CoV-2 Omicron variant infection was confirmed by RT-PCR via daily nasopharyngeal swabs. All eligible patients were divided into those who received Azvudine (Azvudine group) and those who received exclusive supportive treatment (control group).

### Outcomes and definitions

The primary outcome is the time from the first dose of Azvudine (or supportive treatment) to the time when the first negative nucleic acid result was obtained. The nucleic acid test with samples obtained by nasopharyngeal swabs was repeated daily. The secondary outcomes include the nucleic acid conversion rate within the first five days after drug treatment and during the hospitalization, the total course of COVID-19, recurrence of positive nucleic acid results during the first month after hospital discharge, length of hospital stay, and incidence of adverse events. The total course of COVID-19 was defined as the interval between the diagnosis of Omicron variant infection and when the first negative nucleic acid result was obtained. The different clinical categories of COVID-19 were defined according to the Chinese Diagnosis and Treatment Program for Novel Coronavirus Pneumonia (Ninth Edition). The definition of mild COVID-19 was patients with mild clinical symptoms and without signs of pneumonia in imaging, whereas moderate COVID-19 was defined when patients had fever, respiratory symptoms, and signs of pneumonia in imaging.

### Data Collection

Demographic and clinical characteristics, including age, gender, clinical category, risk factors for critical COVID-19, and symptoms at admission, were collected from electronic medical records. Daily cycle threshold of nucleic acid and adverse events during the Azvudine treatment were also collected.

### Statistical Analysis

All data analyses were performed using R software (version 4.1.0). The distribution of continuous variables was examined for normality using the Shapiro-Wilk test. Continuous variables were expressed as median (interquartile range) and analyzed by Mann–Whitney U test. Categorical variables were expressed as absolute numbers (percentage) and compared by Pearson’s chi-square or Fisher exact test as indicated.

Linear regression models were used to investigate the association between Azvudine treatment and time to obtain the first negative nucleic acid test results in unadjusted and adjusted models. Subgroup analyses were performed according to age (≤60 and >60 years old), and clinical categories (asymptomatic and mild-to-moderate COVID-19). Mean differences and 95% CIs were calculated. We also applied the Cox proportional hazards models to assess the relationship between Azvudine treatment and the rate of nucleic acid negative conversion. Variables, including age, gender, group (Azvudine or control group), and clinical categories, were tested by univariable and multivariable analysis. Pearson’s correlation coefficient and the variance inflation factor were used to detect the presence of multicollinearity among variables included in the regression model. Kaplan-Meier methods were used to display curves for time to the first negative nucleic acid test within the first five days after drug treatment. A log-rank test was conducted to compare the time to the first negative nucleic acid test curves of the two groups. A two-tailed p-value of <0.05 was considered significant.

## Results

### Patients enrolment and characteristics

A total of 207 patients were included in the analysis (Figure 1). Among this cohort, 166 patients (80.2%) received Azvudine treatment after admission. The demographic and clinical characteristics of the study patients are summarized in Table 1. Patients in the Azvudine group were older than the control group [29 (7-47) vs. 36 (27-47), P=0.003]. The majority of the study subjects were asymptomatic (79/207, 38.2%) or mild COVID-19 (111/207, 53.6%), and had no risk factors for critical COVID-19 (169/207, 81.6%).

**Figure 1.**
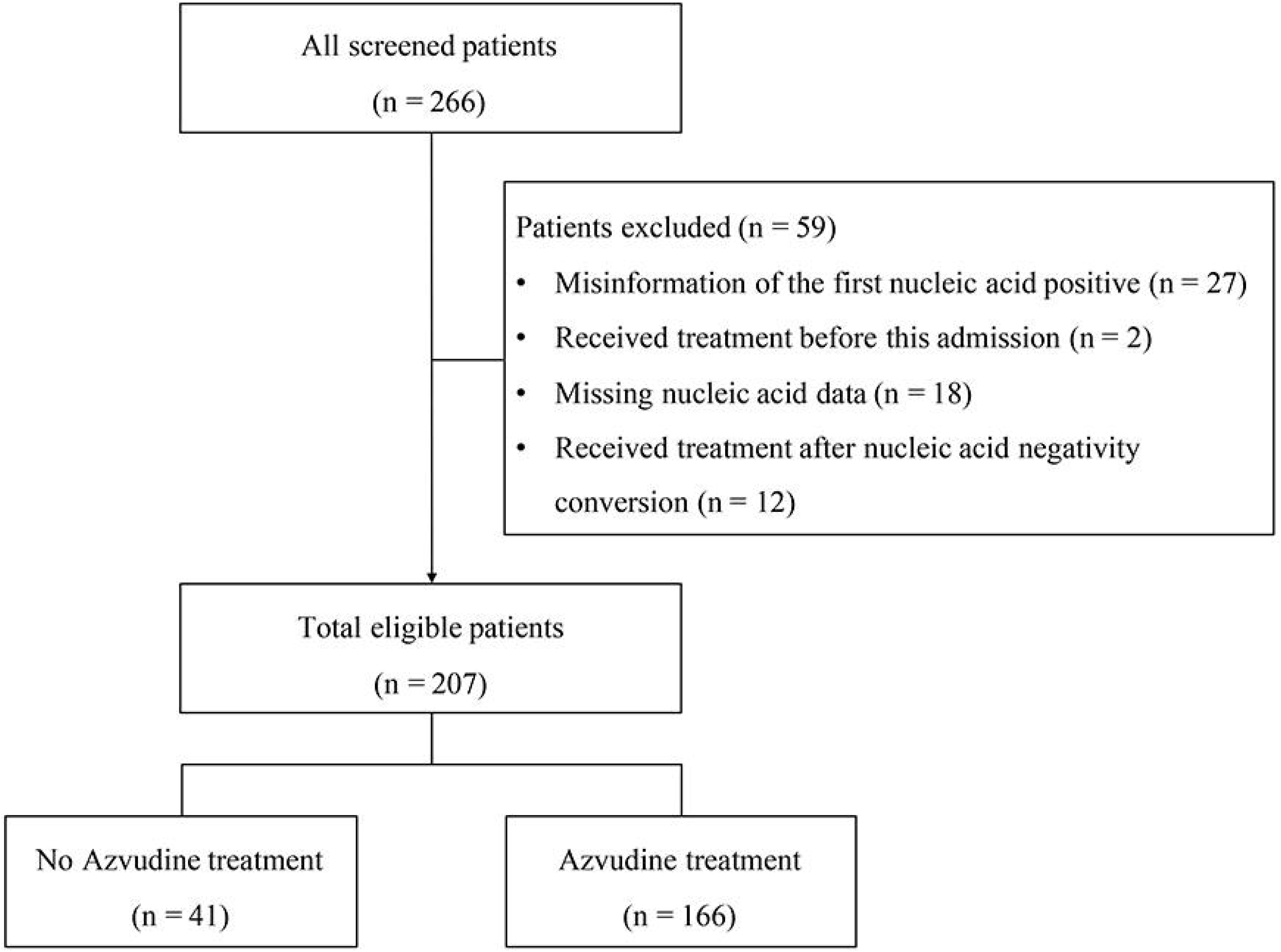
Flowchart of study subjects

**Table 1.**
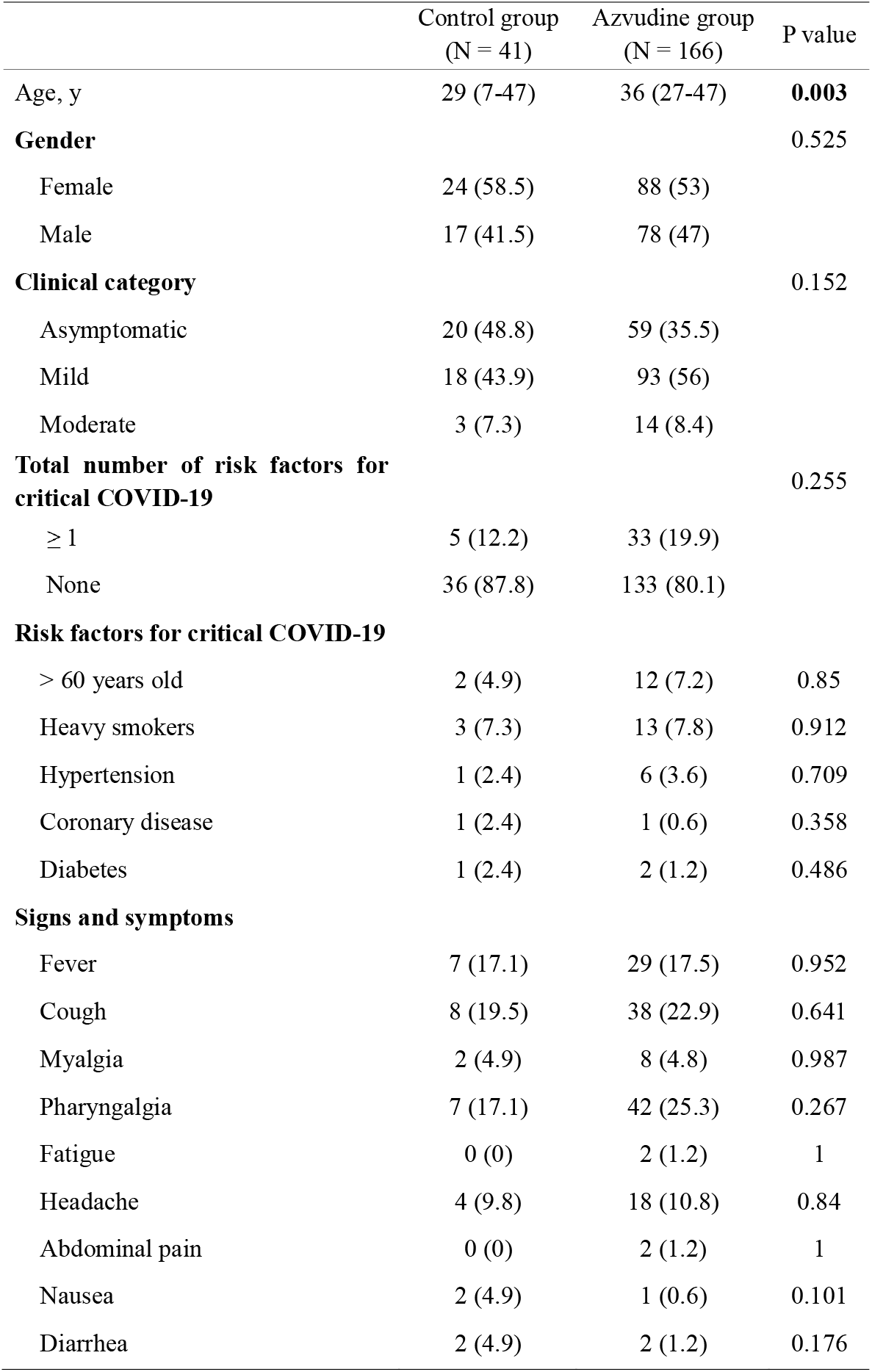
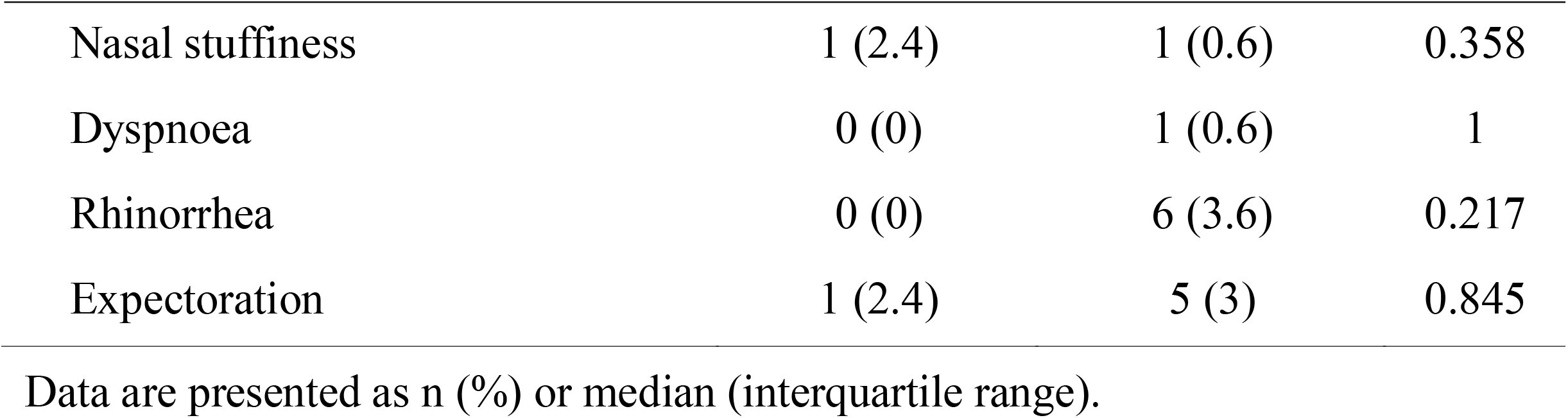
Demographic and clinical characteristics of study patients

The most common signs or symptoms at the onset of COVID-19 were fever (36/207, 17.4%), cough (46/207, 22.2%), and pharyngalgia (49/207, 23.7%). There were no significant differences in gender, clinical categories, risk factors for critical COVID-19, and signs or symptoms between groups.

### Clinical outcomes and adverse events

The clinical outcomes and adverse events of the study patients are shown in Table 2. All patients survived and obtained negative nucleic acid test results before discharge. The median course of SARS-CoV-2 infection and length of hospital stay were 8 (IQR: 7-11) days and 12 (IQR: 9-15) days, respectively. A total of 13 patients had a recurrence of positive nucleic acid test, including four patients in the control group (9.8%) and nine in the Azvudine group (5.4%). Patients receiving Azvudine treatment had a shorter time from initiation of treatment to the first negative nucleic acid test results [5 (IQR: 1-7) vs 6 (IQR: 5-7) days, P = 0.001]. Nine patients had adverse events, and nausea, diarrhea, and vomiting were the most common adverse events in the Azvudine group.

**Table 2.** Clinical outcomes and adverse events of study patients

|  | Control group<br>(N = 41) | Azvadine group<br>(N = 166) | P<br>value |
| --- | --- | --- | --- |
| Proportion of nucleic acid negative conversion | 41 (100) | 166 (100) |  |
| Time from drugs treatment to first nucleic acid-negative conversion | 6 (5-7) | 5 (1-7) | 0.001 |
| Total course of COVID-19 | 8 (7-13) | 9 (7-11) | 0.945 |
| Recurrence of nucleic acid positive | 4 (9.8) | 9 (5.4) | 0.306 |
| Length of hospital stay | 12 (10-14) | 12 (8-15) | 0.793 |
| Proportion of subjects with adverse events | 0 (0) | 9 (5.4) | 0.273 |
| **Adverse events** |  |  |  |
| Nausea | 0 (0) | 6 (3.6) | 0.474 |
| Diarrhea | 0 (0) | 6 (3.6) | 0.474 |
| Vomiting | 0 (0) | 6 (3.6) | 0.474 |
| Stomachache | 0 (0) | 2 (1.2) | 1 |

Data are presented as n (%) or median (interquartile range).

### Association between drugs treatment and first nucleic acid negative conversion

As shown in Table3, Azvudine treatment was associated with reduced time to obtain the first negative nucleic acid test results after adjusting for age and gender [mean difference = −1.658; 95% CI: −2.772 to −0.544, P = 0.0039]. In subgroup analyses, the association remained significant in younger patients (≤60 years old, P = 0.0092), but not in elderly patients (>60 years old, P = 0.0986). Moreover, the efficacy of Azvudine was not impacted by the disease categories (asymptomatic or mild-to-moderate COVID-19).

**Table 3.**
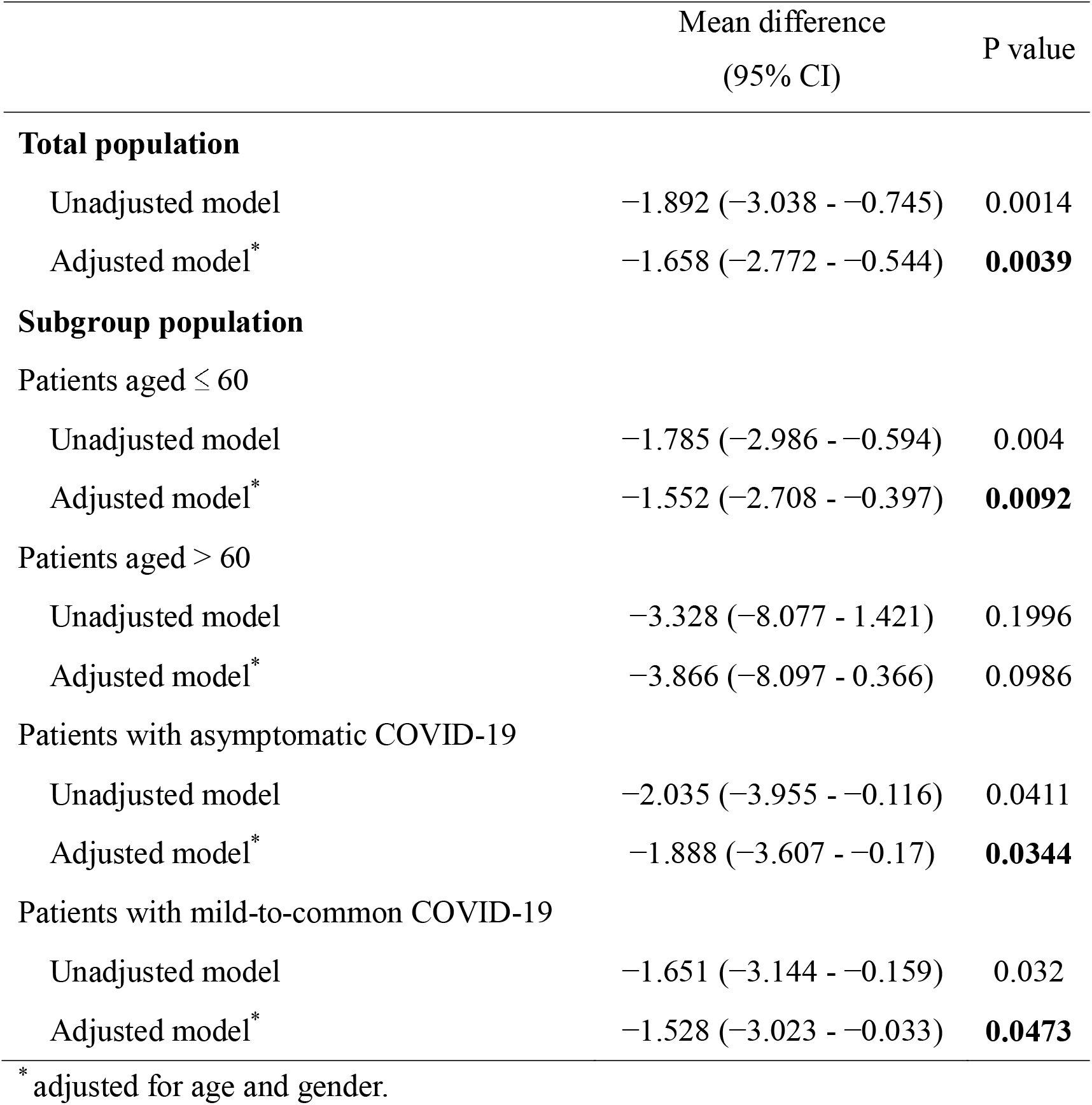
Relationship between Azvudine treatment and time to obtain the first negative nucleic acid test results

|  | Mean difference<br>(95% CI) | P value |
| --- | --- | --- |
| **Total population** |  |  |
| Unadjusted model | -1.892 (-3.038 - -0.745) | 0.0014 |
| Adjusted model* | -1.658 (-2.772 - -0.544) | **0.0039** |
| **Subgroup population** |  |  |
| Patients aged $\leq 60$ | | |
| Unadjusted model | -1.785 (-2.986 - -0.594) | 0.004 |
| Adjusted model* | -1.552 (-2.708 - -0.397) | **0.0092** |
| Patients aged $> 60$ | | |
| Unadjusted model | -3.328 (-8.077 - 1.421) | 0.1996 |
| Adjusted model* | -3.866 (-8.097 - 0.366) | 0.0986 |
| Patients with asymptomatic COVID-19 |  |  |
| Unadjusted model | -2.035 (-3.955 - -0.116) | 0.0411 |
| Adjusted model* | -1.888 (-3.607 - -0.17) | **0.0344** |
| Patients with mild-to-common COVID-19 |  |  |
| Unadjusted model | -1.651 (-3.144 - -0.159) | 0.032 |
| Adjusted model* | -1.528 (-3.023 - -0.033) | **0.0473** |

\* adjusted for age and gender.

The multivariable Cox analysis also revealed that the Azvudine treatment was associated with an increased rate of nucleic acid negative conversion during the hospitalization (HR = 1.461; 95% CI, 1.01 to 2.11, P = 0.044) (Table 4). Additionally, the Kaplan–Meier survival curves illustrated Azvudine treatment could increase the nucleic acid negative conversion rate within the first five days after drug treatment (Figure 2, P_log-rank test_ = 0.001).

**Table 4.** Cox proportional hazards model variables for the rate of nucleic acid negative conversion during hospitalization

|  | Univariable analysis |  | Multivariable analysis |  |
| --- | --- | --- | --- | --- |
|  | Hazard ratio (95% CI) | P value | Hazard ratio (95% CI) | P value |
| Age | 0.999 (0.991-1.007) | 0.849 | 0.997 (0.988-1.006) | 0.518 |
| **Group** |  |  |  |  |
| Control group | Reference |  | Reference |  |
| Azvudine group | 1.407 (0.988-2.004) | 0.058 | 1.461 (1.01-2.113) | **0.044** |
| **Gender** |  |  |  |  |
| Female | Reference |  | Reference |  |
| Male | 1.078 (0.818-1.421) | 0.594 | 1.086 (0.823-1.434) | 0.56 |
| **Clinical category** |  |  |  |  |
| Asymptomatic | Reference |  | Reference |  |
| Mild | 1.004 (0.752-1.341) | 0.978 | 0.969 (0.723-1.299) | 0.833 |
| Moderate | 0.918 (0.543-1.554) | 0.751 | 0.92 (0.535-1.582) | 0.764 |

**Figure 2.**
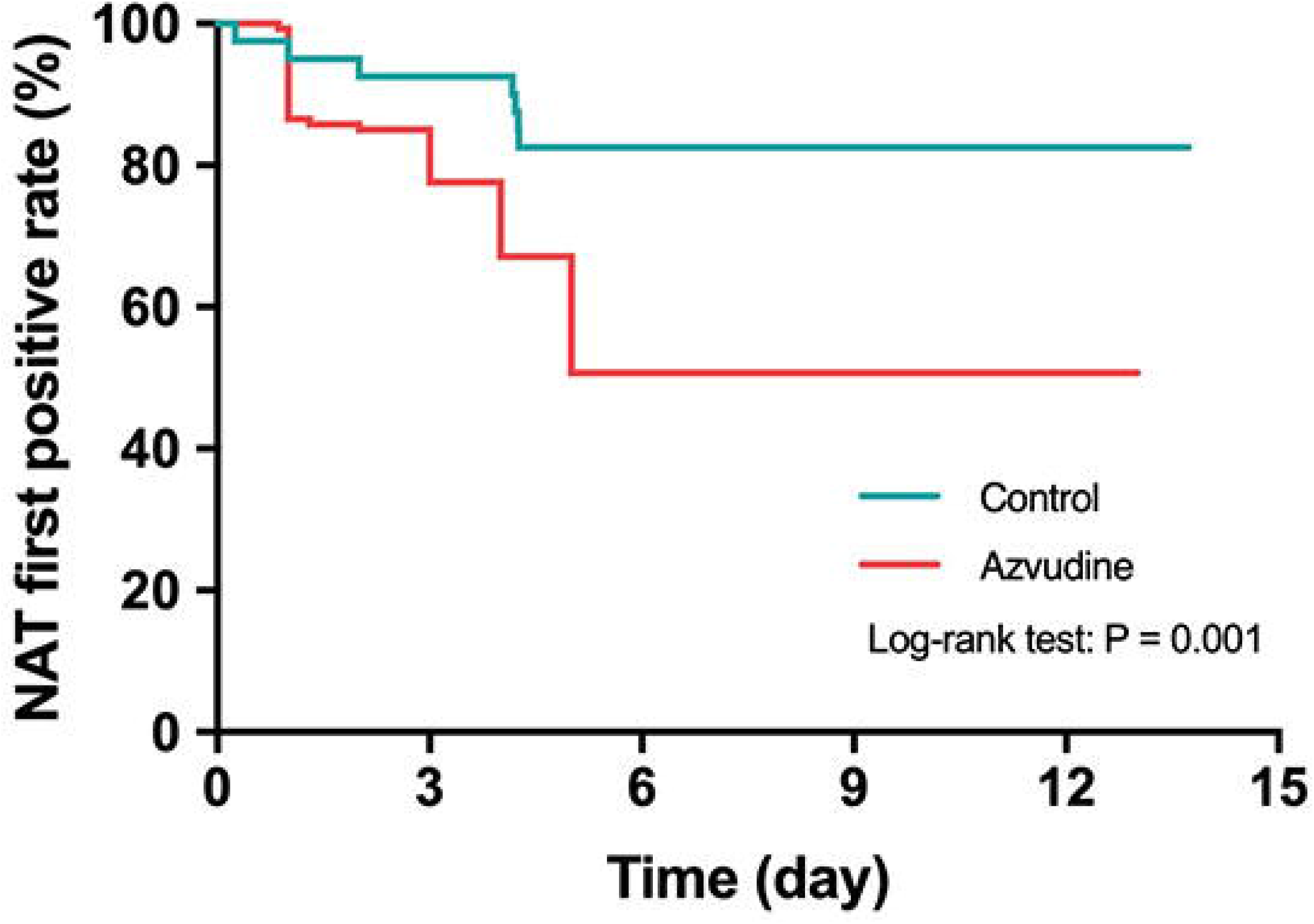
Kaplan–Meier curves of nucleic acid testing negativity within the first five days after drug treatment

## Discussion

This study found that most patients infected by the Omicron variant had asymptomatic or mild COVID-19. Moreover, Azvudine treatment was associated with a reduced time to obtain the first negative nucleic acid test results. To the best of our knowledge, this is the largest study to evaluate the effect of oral Azvudine in patients infected with the SARS-CoV-2 Omicron variant.

Our results are consistent with an observational study that the majority of patients infected with the SARS-CoV-2 Omicron variant are asymptomatic or mild COVID-19 [11], suggesting a reduced pathogenicity of Omicron variants [12, 13]. Nevertheless, the unique biological characteristics of Omicron variants increase the transmissibility, particularly in asymptomatic COVID-19 patients [14]. These patients may recover faster if they receive timely antiviral agents. However, there is a lack of effective antiviral agents, which may accelerate the spread of the pandemic and account for a large cumulative expense of medical resources.

Azvudine is a promising antiviral agent for COVID-19 treatment [15]. A previous animal study showed that oral Azvudine (0.07 mg/kg, qd) could reduce the viral load and alleviate inflammation response or organ damage [16]. Recently, a pilot study recruited 20 patients with mild and moderate COVID-19 and randomly assigned them to receive Azvudine (n = 10) or standard antiviral treatment (n =10). The results showed that oral Azvudine tablets could shorten the viral shedding time compared with standard antiviral treatment [10], which is similar to our study. Additionally, a randomized, single-arm clinical trial showed that oral Azvudine (5 mg, qd) could achieve 100% viral nucleic acid conversion in 3.29 ± 2.22 days (IQR: 1–9 days) and 100% hospital discharge in 9.00 ± 4.93 days (IQR: 2–25 days) [16].

On the contrary, other antiviral agents did not exhibit satisfying results. Remdesivir can directly inhibit the activity of SARS-CoV-2 in vitro [17], but a recent report about the compassionate use of remdesivir for patients with severe COVID-19 showed that clinical improvement was only observed in 68% of patients [18]. Nirmatrelvir is approved by the Food and Drug Administration for the treatment of mild-to-moderate COVID-19 in preventing severe progression, and a large retrospective cohort study showed that Nirmatrelvir only reduced the rate of severe COVID-19 in patients aged ≥ 65 years old [19]. Our subgroup analysis showed that Azvudine might also be effective in patients under 60 years old.

The strength of this study is that this is the largest study to investigate the effect of Azvudine treatment in patients infected with the SARS-CoV-2 Omicron variant. However, several limitations should be addressed. First, although several attempts were made to control the unbalance of demographic and clinical characteristics between groups, unmeasured confounders cannot be adjusted. Moreover, we did not collect data regarding routine laboratory tests and other variables of clinical improvements. Additionally, owing to the retrospective nature of this study, a causal relationship cannot be inferred, and the results should be interpreted with caution.

## Conclusion

Our study showed that Azvudine treatment was associated with reduced time to obtain the first negative nucleic acid test results. Larger randomized clinical trials are needed to assess the impact of oral Azvudine tablets on clinical improvement in patients infected with the SARS-CoV-2 Omicron variant or future variants.

## Data Availability

The datasets are not publicly available due to privacy but are available from the corresponding author on reasonable request.

## Declarations

## Acknowledgment

None.

## Authors’ contributions

WL, GQW equally contributed to the conception and design of the research. WMC, HGX, LH, RY and CQP performed the research and analyzed the data. WMC, HGX and LH wrote the manuscript and interpretation of data. All authors critically revised the manuscript, agree to be fully accountable for ensuring the integrity and accuracy of the work, and read and approved the final manuscript.

## Ethics statement

This study was approved by the institutional review board of the First Affiliated Hospital of Bengbu Medical College with a waiver of consent for the the retrospective observational design (No. 2020KY112, 2020KY016).

